# Increasing household diet diversity and food security in rural Rwanda using small-scale nutrition-sensitive agriculture: A community-level proof of concept study

**DOI:** 10.1101/2022.09.27.22280437

**Authors:** Brittney C. Sly, Tiffany L. Weir, Christopher L. Melby, Leslie Cunningham-Sabo, Stephen J. Leisz, Valerie J. Stull

## Abstract

**Objectives:** In Rwanda, rates of malnutrition remain high in rural areas where residents consume a primarily starch-based, low variety diet. Nutrition-sensitive agricultural interventions using kitchen gardens have been effective in addressing low diet diversity in similar populations. This study’s objective was to develop a kitchen garden and nutrition education intervention aimed at sustainably increasing diet diversity and food security at the household-level.

**Design:** A mixed methods community-level study, with a sixteen-week nutrition-sensitive agricultural intervention including nutrition education was conducted. Household diet diversity scores and household hunger scores were calculated at baseline, post-intervention and one-year follow-up.

**Setting:** The intervention was conducted in a rural Rwandan community in the Northern Province.

**Participants:** Stratified purposeful sampling techniques were used to select women participants representing forty-two households.

**Results:** Household diet diversity scores increased over time from pre-intervention to six months post-intervention and one-year post-intervention. The magnitude of the change was similar in all stratified groups (2.3x at 6 months and 2.9x at 1 year). Households whose main source of income was working for other farmers, reported a significantly lower diet diversity score than those households receiving income from sources [t(40) = -2.108, p=0.041]. Among those households not consuming protein and vitamin-A rich food groups at baseline, all reported consuming foods from these food groups post-intervention. There were no significant changes in household hunger scores.

**Conclusions:** Collaborative community-based nutrition-sensitive agricultural interventions using kitchen gardens, can increase household diet diversity, which may encourage sustained change in dietary patterns for nutritional adequacy in low-income rural Rwandan populations.

## Introduction

In 1994, Rwanda experienced a devastating genocide that decimated the population, infrastructure, and the economy. Since then, Rwanda has made admirable progress in rebuilding its economy and infrastructure, while improving living standards for its population of 12.5 million people. However, a large proportion of the population still faces pervasive poverty, malnutrition, and food insecurity. Recent surveys indicate that 39.1% of the population lives in poverty, concentrated in rural areas where 83% of people reside.^1^ Over 70% of the population engages in the agricultural sector for income, supplying 90% of the country’s food needs, with most practicing subsistence farming.^1^ Despite national efforts targeted at rural residents, any improvement in rates of malnutrition, particularly among women and children, have stagnated, attributed to a lack of improvement in household food security and adequate diet consumption.^2,3^ In 2015, Rwanda reported 16.8% of households remaining food insecure, predominantly in rural areas where agriculture is the main income source.^3^ Additionally, 23.8% of households were consuming an inadequate diet, meaning the variety of foods consumed as determined over a seven day period was less than optimal to ensure sufficient nutritional balance.^3^ These data corroborate the estimated 48% of children age 6-59 months who are vitamin A deficient, and the 33% of children under the age of 5 who are classified as stunted, with the greatest proportion represented by children living in rural areas.^4,5^ Approximately 13% of women aged 15-49 years of age are anemic, with a higher prevalence among pregnant women living in poorer households.^4^ In the Burera District, a rural area located in the Northern Province of Rwanda, nutrition-related indicators show a more severe situation with approximately 30% of households reporting food insecurity and 45-63% reporting inadequate food consumption, based on food consumption scores.^3^ The prevalence of childhood stunting (43%) is higher than the national rate (38%), and the prevalence of anemia in women of reproductive age is 14%.^4^

Despite most rural residents engaging in agriculture, prior work with this population has shown a general lack of knowledge regarding small-scale vegetable production and nutritious food choices and behaviors, particularly regarding diet diversity. Traditionally, the woman head of household prepares daily meals from available local food, gathered from cash crop stores or local markets, resulting in a primarily starch-based, low variety diet. Small-scale fruit and vegetable production via kitchen gardens has been an effective solution for addressing these issues in similar populations.^6–10^ According to a 2016 review by Pandey *et al*., of 25 studies examined, interventions that increased household crop diversity and livestock ownership also increased household-level diet diversity, child dietary diversity, and the consumption of nutrient-dense animal foods, fruit and vegetables, as well as increased micronutrient intake.^11^ Another review concluded that projects providing agricultural resources, while also investing in human capital through nutrition education and gender considerations, had the greatest likelihood of effecting positive changes in nutrition.^12^ Specifically, small-scale fruit and vegetable production via kitchen garden projects were identified as nutrition-sensitive agriculture interventions having the highest success rate due to their ease of adoptability, investment in human capital, and women’s empowerment aspects.^12^ Therefore, implementing education and support for kitchen gardening, targeted towards rural Rwandan women, has the potential to combat malnutrition by increasing household diet diversity while also providing a stable supply of food for the household.

Using participatory action research approaches and peer-training methods, an interventional model was developed aimed at educating and empowering women to expand on their existing agricultural and nutrition knowledge with the primary goal of increasing household food security and diet diversity through kitchen-gardening. There were two specific aims: 1) to enhance household dietary diversity by increasing consumption of a variety of home-grown fruits and vegetables, and 2) to improve household food security by providing a more consistent food source via kitchen gardens. It was hypothesized that this intervention would result in changes in dietary patterns consistent with improved diet quality and increased food security. Sustainable increases in these parameters could ultimately remediate and prevent malnutrition. Finally, this research could provide a collaborative model to be used as a basis for program design and evaluation with similar populations and communities that face malnutrition and food insecurity at a regional and global scale.

## Materials and Methods

This study was conducted in a rural community within the Northern Province of Rwanda. The research was done in collaboration with a U.S. based non-governmental organization (NGO) that has worked within the community since 2006. Prior to the start of the study, a comprehensive exploratory assessment was conducted with a sample of the intended study population. Extensive meetings were also held to discuss the intervention and work with community leaders to establish the intended methods of the study and drive the development of the intervention materials.

### Study Design

#### Sample Selection and Setting

Study participants were recruited from the Cyanika area located in the Burera District in northern Rwanda. Women were selected for this intervention study because of their traditional role as decision makers regarding food for the household. Past research has identified women as an ideal conduit for malnutrition remediation interventions in rural poor agricultural-based populations throughout sub-Saharan Africa.^9,13,14^ Forty-two non-pregnant women >18 years of age and considered the woman head of household, were selected by community liaisons using a stratified purposeful sampling method. Participants were chosen based on the geographic location of their permanent residence, their perceived need for assistance, and their willingness to train others in the future. Each participant was then placed in a stratum for the duration of the study. Purposeful sampling allowed for a limited number of cases to reach the goal of acquiring in-depth analysis that could guide the investigator understand the central problem under study.^15^ This method was appropriate because of the homogeneity of the groups and the research goal of examining variation in key variables.^16^ Randomized sampling was infeasible as the long-term goal of the intervention was to provide sustained change and generate information spread within a specific community. Therefore, by allowing community liaisons, who were respected members of the community, to recruit and choose participants based upon the study criteria, it allowed for greater community buy-in, which was considered essential to the success of the intervention. Subjects were recruited from each of the six, government-established geographically specific community groups, referred to as *cells*. Each cell contained seven participants, and a leader for each cell was chosen internally by the group. The initial forty-two participants were referred to as *project ambassadors* to foster empowerment and emphasize their future role as trainers. This study was approved by the Institutional Review Board at Colorado State University (ID: 19-9040H). All participants were fully informed of the research and written informed consent was obtained. The study was conducted according to the guidelines provided in the Declaration of Helsinki.

#### Intervention

From January through May 2019, each cell group participated in a 16-week intervention that included lecture-based trainings, hands-on farming activities and socialization. Members of each cell met weekly for approximately 2 hours in their separate cell groups with project coordinators and subject-matter community experts, such as a *Community Nutrition Health Worker* and an *Agronomist*, at the group leader’s home, known as the *Demonstration Site*. The time commitment was kept to what was considered reasonable by participants, but still allowed for ample return on their commitment through increased knowledge and home-grown vegetable crops. Materials and curriculum specific to this study were developed to educate participants and provide specific learning topics on a weekly basis (**Table 1**.).

**Table 1.** Weekly intervention curriculum and learning topics.

| Week | Learning Topic |
| --- | --- |
| 1 | Introduction to basic nutrition and home gardens |
| 2 | Trench garden design |
| 3 | Keyhole garden design |
| 4 | Work week |
| 5 | Water management and the nutritional needs of various household members part 1 |
| 6 | Composting and different nutrients in food |
| 7 | Work week |
| 8 | Pest and weed management and the nutritional needs of various household members part 2 |
| 9 | Seed saving and household meal planning |
| 10 | Work week |
| 11 | Perennial crops, fruit trees and nutritional deficiencies |
| 12 | Food safety and preservation and ensuring household food security |
| 13 | Cooking and planning balanced meals |
| 14 | Work week |
| 15 | Question and answer session |
| 16 | Large group reflection and celebration |

Participants created keyhole-style and raised-bed gardens at their homes, as well as compost piles and rainwater catchment systems for irrigation. They also learned about how to prepare balanced meals for all members of their household using the fruits and vegetables they grew in their kitchen garden. Resources were provided throughout the intervention, including workbooks and notepads, gardening tools, seeds, and construction materials – Some resources were intended for personal-use and others to be shared. Due to the varied education and literacy levels, materials included primarily pictures and illustrations.

### Data Collection

Both qualitative and quantitative data were collected at three time-points during the study, selected to capture seasonal variations in food availability: 1) baseline data was collected in November 2018 during a time this region experiences fairly constant weather patterns with occasional rainfall and mild temperatures, 2) six-months post intervention in September 2019 during what is considered their ‘sunny’ season, characterized by low rainfall, limited crop growth and historically low food security and diet diversity, and 3) one-year post intervention in June and July 2020 in order to evaluate the sustainability of intervention goals.^4^ Qualitative data acquired via focus groups and interviews are discussed elsewhere^.17^

Semi-structured interviews were used to collect data on diet diversity and food security. The lead researcher (BS) conducted interviews with the assistance of a local translator. Interview questions and responses were translated in real-time from English to Kinyarwanda and back by a trained translator. The translator was a university student and local member of the community, who completed training in research ethics along with conceptual training with the lead researcher by way of a bilingual individual not involved in the study. From these instructional exercises, the research team determined the interviewer and translator to be able to collect accurate data in accordance with the study protocol.

#### Socio-demographics

Sociodemographic details were collected from interviewer-administrated questionnaires completed during semi-structured interviews. Data were obtained concerning household size and occupants, marital status, sources of income, sources for obtaining food, and whether they engaged in cash-crop agriculture.

#### Dietary Assessment

Dietary assessment was performed using a 24-hour open recall administered by a trained dietitian to estimate the ‘usual’ intake of the household at each of the three time-points using the woman head of household as the proxy. Participants were asked to include all foods eaten by all household members, including meals, snacks, and foods eaten within and outside the home.

Although using multiple 24-hour recalls over several days is considered the best reference for assessing diet diversity, evidence from prior research indicate that using a single 24-hour recall in rural sub-Saharan African populations is sufficient to predict nutritional status and regular dietary intake as compared to a standard three-day recall.^18^

#### Household Diet Diversity Score

Dietary information gathered from respondents was applied to a diet diversity equation as done in previous studies with similar populations, calculated by summing the number of different food groups consumed by individuals in a household over the 24-hour recall period.^9^ Information recorded from the 24-hour recalls was used to determine a household diet diversity score (HDDS) based upon adaptation of the Food and Agriculture Organization (FAO) tool to include regional and cultural-specific foods.^19^ For the purpose of discussion, protein foods are defined as foods from groups: *Organ and Flesh Meat, Eggs, Fish and Seafood, Legumes, Nuts and Seeds*, and *Milk and Milk Products*. HDDS is a qualitative measure that can serve as a proxy for the nutrient adequacy of the diet of individuals while also reflecting household access to a diversity of foods.^9,19,20^ The HDDS is calculated based on the number of food groups consumed out of twelve possible food groups (**Table 2**).^19^ A second coder not involved with the collection of the original data, independently applied the dietary assessment data from seventeen participants, representing 15% of the total sample, to the diet diversity calculator to check inter-rater reliability. 100% agreement was established between the coders.

**Table 2.** A list of the 12 food groups and the foods contained within each group according to the FAO, adapted for regional and cultural-specific foods. Specific food groups of nutritional interest are bolded.

| **Food Groups** | **Foods Within the Food Group** |
| --- | --- |
| Cereals | maize, rice, wheat, sorghum, millet, or any other grains or foods made from these (e.g., bread, noodles, ugali) |
| White Roots and Tubers | white potatoes, white yams, or white cassava |
| Vegetables | **<u>Vitamin A Rich Vegetables and Tubers:</u> pumpkin, carrot, squash, or sweet potatoes that are orange inside, ibihaza, tomato**<br>**<u>Dark Leafy Green Vegetables:</u> dark green leafy vegetables including spinach, kale and wild forms and/or locally available vitamin-A rich leaves such as from amaranth (dodo), cassava (isombe)**<br><u>Other Vegetables:</u> any other vegetables (e.g., tomato, onion, eggplant, cabbage, onion, lettuce, celery) |
| Fruits | **<u>Vitamin A Rich Fruits:</u> mango, cantaloupe, apricot, papaya, peach, and 100% fruit juice made from these fruits**<br><u>Other Fruits:</u> matoke (banana variety), passion fruit, avocado, plantain, pineapple, and other fruits, to include 100% fruit juices made from these fruits |
| Organ and Flesh Meats | **liver, kidney, heart, or other organ meats, and beef, pork, lamb, goat, rabbit, chicken, duck, other birds, insects and wild game** |
| Eggs | **eggs from chicken, duck, or guinea fowl** |
| Fish and Seafood | **fresh or dried fish and seafood** |
| Legumes, Nuts and Seeds | **dried beans, dried peans, lentils, nuts, seeds, or foods made from these foods (e.g., peanut butter, ikinyiga)** |
| Milk and Milk Products | **milk, cheese, yogurt, or other milk products** |
| Oils and Fats | oil, butter, margarine added to food or used for cooking |
| Sweets | sugar, honey, sweetened beverages, sugary foods such as candies, cookies, cakes, sweet bread foods |
| Spices, Condiments and Beverages | spices (salt and black pepper), condiments (ketchup sauce, mayonnaise), coffee, tea, alcoholic beverages |

#### Food Security Evaluation

Household food security was evaluated during interviews at all three data collection time-points. The Household Hunger Scale developed by the Food And Nutrition Technical Assistance (FANTA) Project was used because of its validity depicting comparable indicators across cultures and within food insecure populations.^21^ Measurements of household hunger were used as a proxy for household food security, through a series of questions used to determine a score for each household, with a higher score equating to more hunger or greater food insecurity.^22^ Three questions were asked during each interview to tabulate a Household Hunger Score (HHS) that represented a household hunger category.^22^ A second coder not involved with the collection of the original data, independently calculated the HHS from recorded interview data for seventeen participants, representing 15% of the total sample, to check inter-rater reliability. 100% agreement was established between the coders.

### Data Analysis

The Statistical Package for Social Sciences software version 26 (SPSS Inc., Chicago, IL, USA) and GraphPad Prism software version 8 (GraphPad Software, San Diego, CA, USA) were used to conduct all statistical analyses. Descriptive data was depicted as means and standard deviations, or percentages. The percentages of households consuming each of the different food groups according to HDDS were determined. A linear mixed-effects model was used to analyze for changes in HDDS and HHS across all time points depicted as means with 95% confidence intervals. ANOVA using a post-hoc Tukey’s test was used to analyze between subject variation, and Pearson’s correlation coefficient was used to determine potential confounders attributing to individual variability in HDDS and HHS, which were then further analyzed using independent *t-* tests. For all statistical analysis, a P value of ≤ 0.05 was accepted as significant.

## Results

Among the forty-two participants, representing their respective households, (mean [SD] age, 41.9 [12.3] years), the majority were married, and a smaller percentage were widowed, separated, or divorced at the time of the study (**Table 3**.). The average number of people living in each household was six, with 66.7% of households having at least one child under the age of five years. Like other rural populations in Rwanda, many participants reported growing staple food crops such as potatoes, maize, beans, sorghum, and sweet potatoes that their household consumed and/or sold for income. The main income source for participants was working in agriculture for other local farmers (66.7%), with 11.8% reporting no source of income. A smaller proportion of households obtained income from a household member working as a retailer of goods at local markets, having formal employment, such as teacher, hair stylist, pastor, or having other sources of income such as charitable gifts from the church or family members.

**Table 3.** Participant characteristics, Cyanika Rwanda, Summer 2020 (n=42).

|  |  |  |
| --- | --- | --- |
| **Age (years), mean (SD)** |  | **41.9 (12.3)** |
| **Marital status, percentage** |  |  |
| **Married** |  | **81.0%** |
| **Widowed** |  | **16.7%** |
| **Separated/Divorced** |  | **2.4%** |
| **Number of people in household, mean (SD)** |  | **6.0 (2.2)** |
| **Number of children in household under the age of 5, mean (SD)** |  | **1.0 (0.9)** |
| **Grow staple crops (potatoes, maize, beans, sorghum and sweet potatoes), percentage** |  | **62%** |
| **Main income source, percentage** |  |  |
| **Retailer** |  | **11.9%** |
| **Work for Other Farmers** |  | **66.7%** |
| **Employed** |  | **4.8%** |
| **Other** |  | **4.8%** |
| **No Income Source** |  | **11.8%** |

At baseline, the average household diet diversity score (mean [SD]) was reported as 2.59 [1.3], with slight variations notably seen amongst the different cell groups. At baseline, prior to the intervention, the three food groups consumed by most households were *White Roots* (55%) in the form of Irish potatoes, *Legumes, Nuts, Seeds* (58%) in the form of dried beans, and *Vegetables* (67%) in the form of leaves from cassava plants, locally known as isombe, or leaves from bean plants (**Figure 1**.). During the data collection period six months post-intervention, increased consumption was reported for all food groups, except for foods included in the *Organ & Flesh Meats, Eggs*, and *Fish* groups. Data collected one-year post-intervention showed a continued increase in the number of all food groups consumed. Most notably, foods in the *Cereals, Vegetables, Spices*, and *Legumes, Nuts, Seeds* food groups were consumed by over 90% of participants.

**Figure 1.**
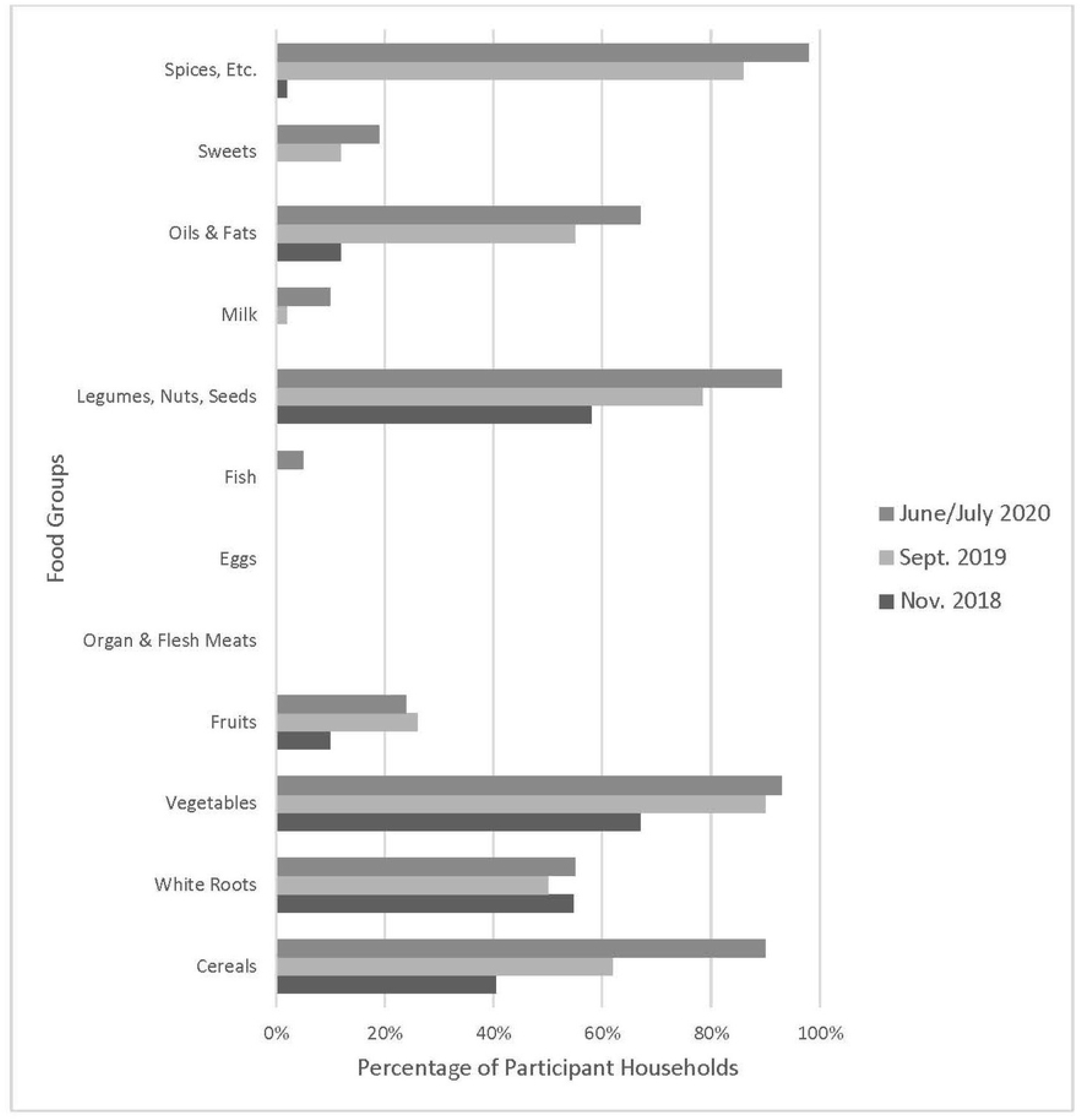
Proportion of participant households consuming foods from each food group during each time point, Cyanika, Rwanda.

Amongst all participant households, the average HDDS significantly increased over time (mean [SD] from pre-intervention (x=2.59 food groups [1.3]), to six months post-intervention (x=4.85 [1.6]) and continued to increase one-year post-intervention (5.55 [1.3]) (**Figure 2**.). The HDDS were on average 2.3 times greater six months post-intervention than pre-intervention, and 2.9 times greater one-year post intervention than pre-intervention, showing a consistent increase over time. These data also indicate that during the ‘sunny’ season, when the availability of food is historically scarcer leading to inadequate and unbalanced dietary patterns, increased household diet diversity was observed.

**Figure 2.**
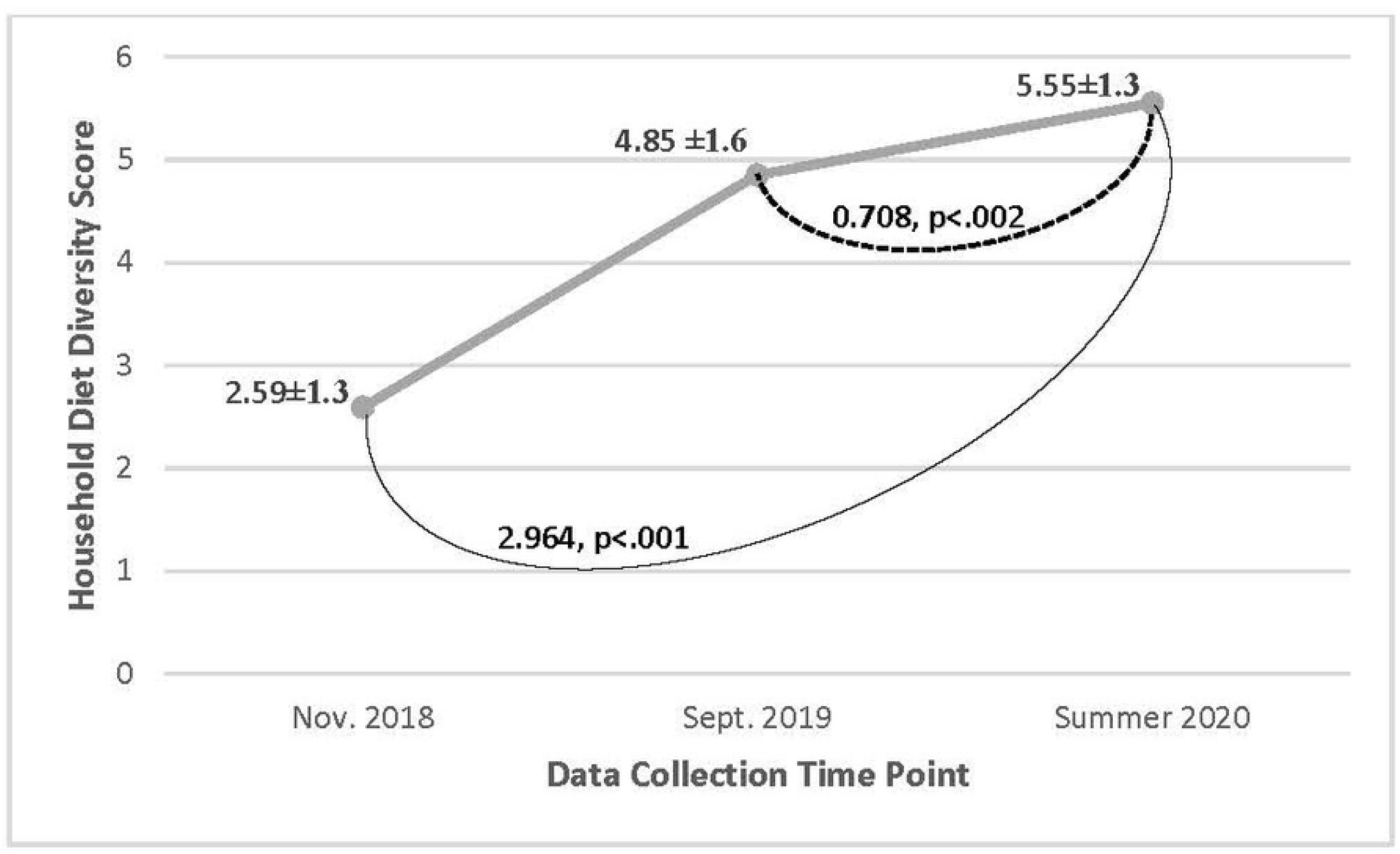
Changes in household diet diversity scores over time for all participant households on a scale of 0-12, Cyanika, Rwanda. Linear mixed effects statistical analysis of the mean household diet diversity scores comparing for changes across the different time points.

When divided into cell groups, the average HDDS continued to increase amongst most participant households in most cell groups (**Figure 3**.). This sustained increased in HDDS was also seen when HDDS was analyzed for each cell group over time. There was a statistically significant effect on HDDS for cell group [F(5, 36) = 3.115, p=0.019], as well as for time for each cell group [F(1.998, 63.92) = 70.70, p<0.001]. This indicates that the magnitude of the change over time was similar in all groups.

**Figure 3.**
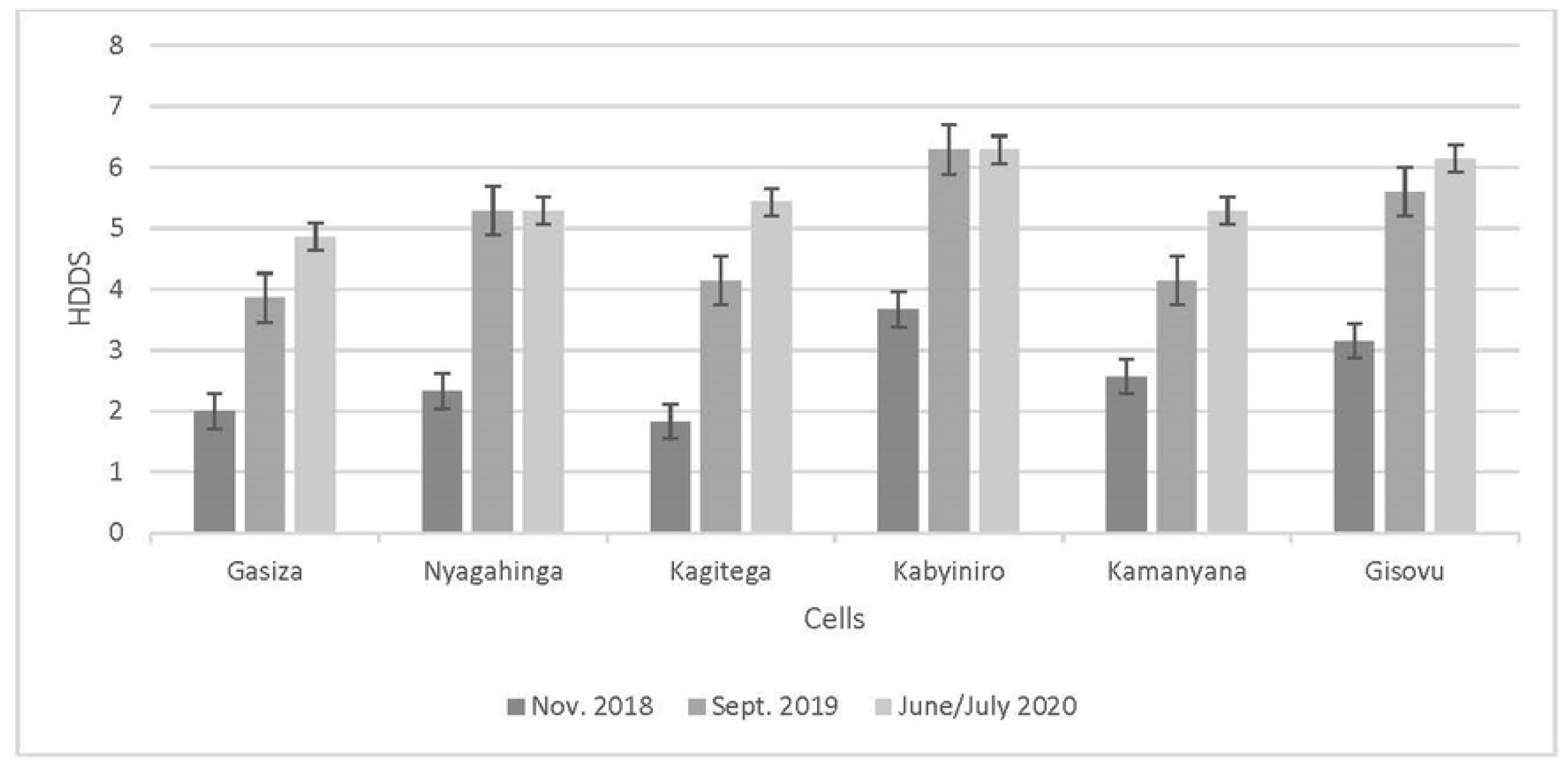
Average household diet diversity scores (HDDS) for participant households by cell group for each time point, Cyanika, Rwanda.

Among households whose main source of income came from working for other farmers, HDDS scores were significantly lower [*t*(40) = -2.108, p=0.041] than for those households with other sources of income (**Figure 4**.). In addition, households whose main source of income came from an employed household member, reported significantly higher HDDS [*t*(39) = 12.940, p<0.001], than those households with other sources of income.

**Figure 4.**
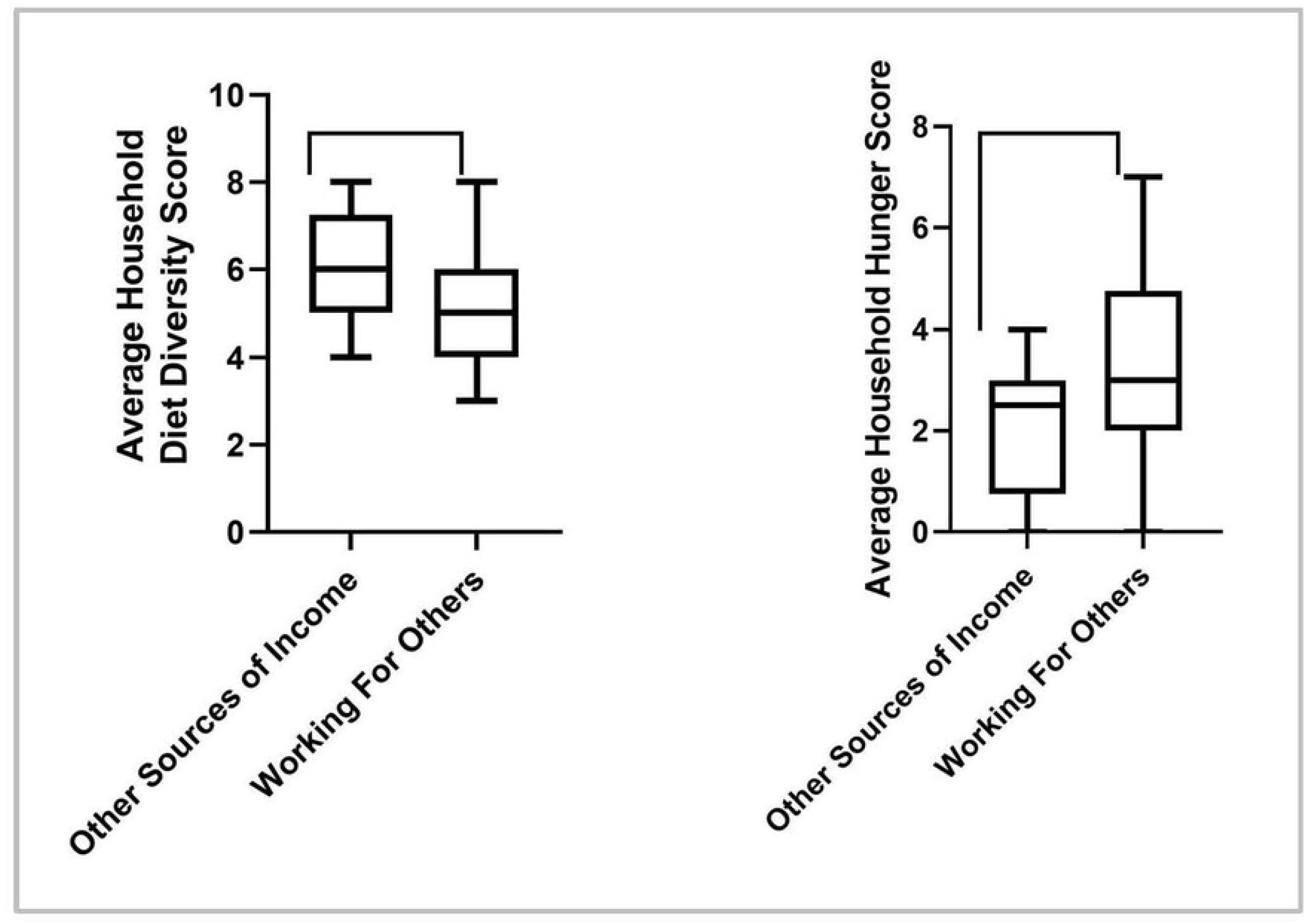
Independent t test analysis showing the negative association between average household diet diversity scores and household hunger scores for women participants, Cyanika, Rwanda, in relation to the main source of household income coming from working for other farmers. The higher the household hunger score, the more food insecure a household.

To determine if the changes in household diet diversity contributed to improved nutritional adequacy, the proportion of participants consuming vitamin A-rich foods and protein foods by cell group at the three time points was examined (**Figure 5a**. and **Figure 5b**.). Among those participants not consuming protein foods and vitamin A-rich vegetables at baseline, after the intervention, all reported consuming foods from these food groups (as defined in Table 2.). In addition, a large majority (83%) of the participants growing vitamin A-rich vegetables in their kitchen gardens, also consumed these vegetables. Thirty-one percent of participants who were consuming ‘other’ vegetables, were also growing them. However, 42% of participants who were growing ‘other’ vegetables, were not consuming them. The vegetables grown at the highest rate were amaranth leaves, a dark green leafy vegetable known locally as dodo (57%), onions (55%), green cabbage (52%), beets (50%), and carrots (43%).

**Figure 5a.**
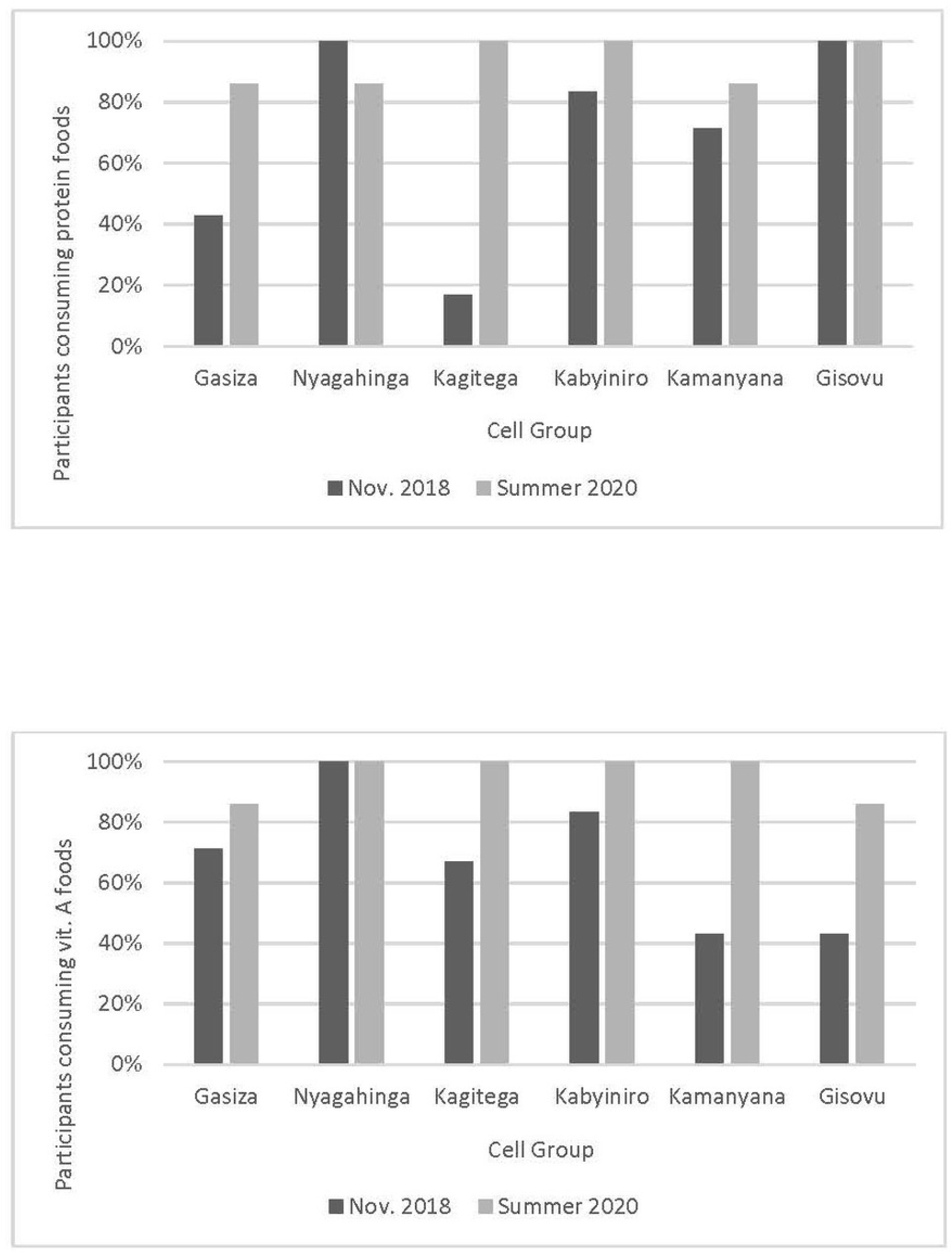
Proportion of participant households in each cell group consuming protein foods pre- and post-intervention, Cyanika, Rwanda. **Figure 5b**. Proportion of participant households in each cell group consuming vitamin A-rich foods pre- and post-intervention, Cyanika, Rwanda.

The results for household food security are represented in Figure 6. and 7. There were no significant changes in HHS scores for any participants across all timepoints, or when divided into cell groups. There were also no significant differences in HHS when examining scores amongst cell groups. Additionally, no relationship was found between changes in HDDS and observed changes in HHS. It was observed however, that households whose main source of income was working for other farmers, reported a significantly higher HHS *t*(40) = -2.090, p=0.043, than those households with other sources of income, indicating greater food insecurity in the former. In addition, households whose main source of income came from an employed household member, reported significantly lower HHS *t*(40) = -2.017, p=0.05, than those households with other sources of income, indicating lower food insecurity.

**Figure 6.**
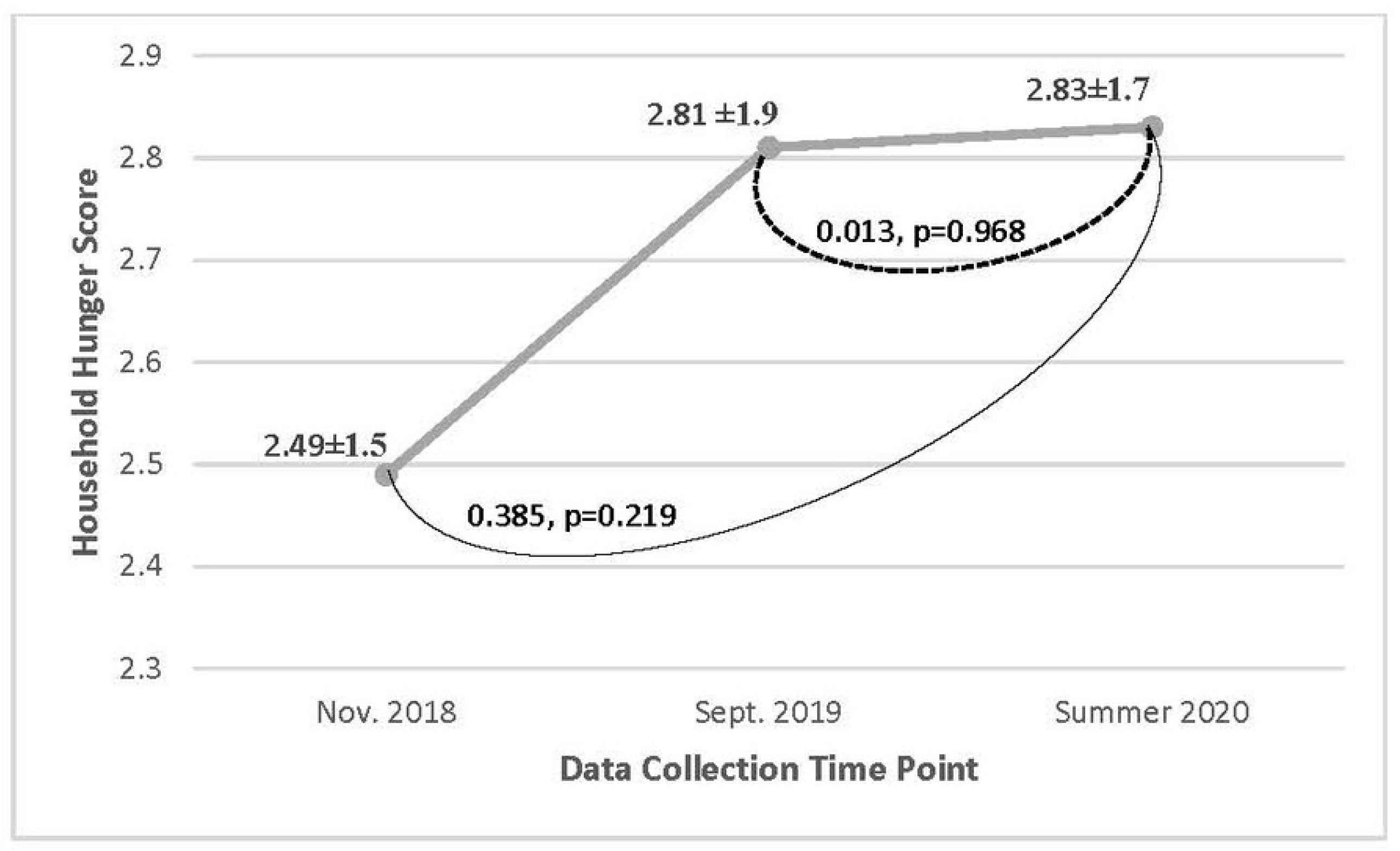
Changes in household hunger scores over time for all participant households on a scale of 0-6, Cyanika, Rwanda. Linear mixed effects statistical analysis of the mean household hunger score comparing for changes across the different time points. Results were non-significant.

**Figure 7.**
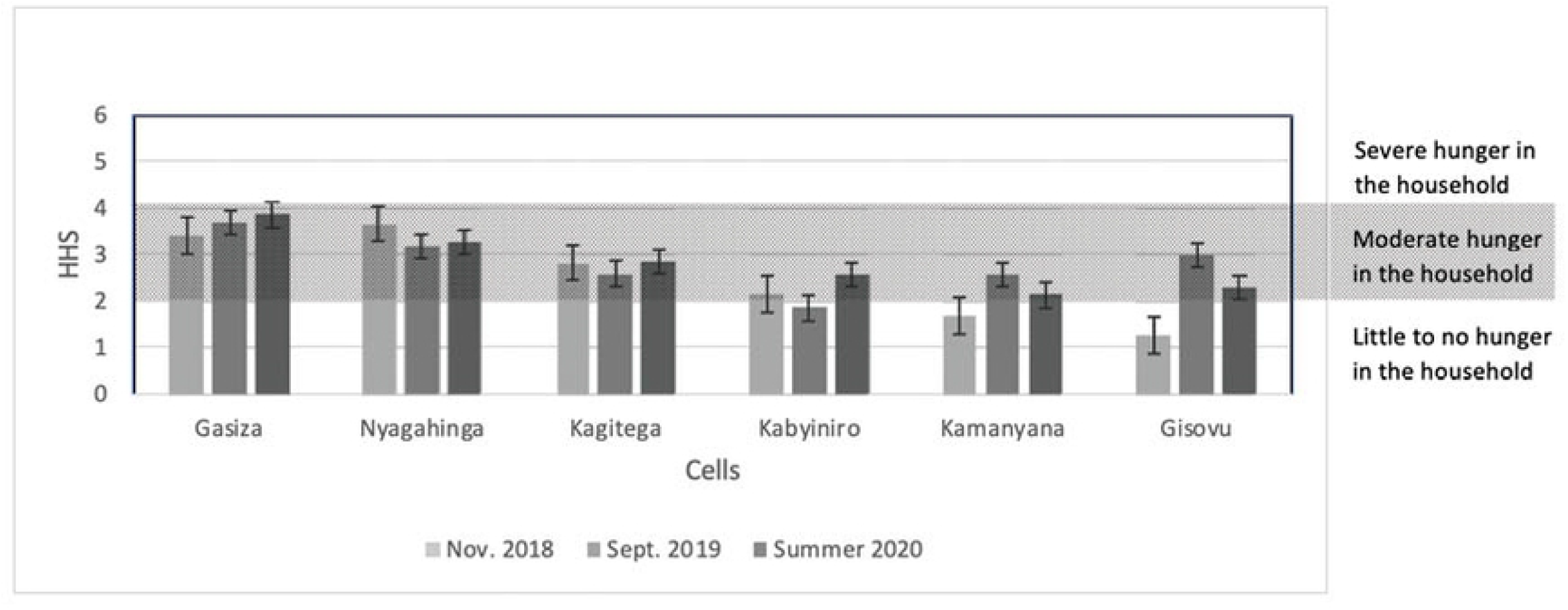
Average household hunger scores (HHS) for participant households by cell group for each time point and their corresponding hunger category, Cyanika, Rwanda, as determined by the Food and Nutrition Technical Assistance project Household Hunger Score (HHS) Tool (22).

## Discussion

It was hypothesized that the nutrition-sensitive kitchen garden intervention would result in a sustained increase in household diet diversity and food security, allowing for improvement in dietary patterns aimed at reducing malnutrition. It was observed that the average household diet diversity increased six-months after the intervention, during the sunny season, as well as one-year after the intervention, showing sustained change. However, the intervention did not result in significant changes in household food security.

### Changes in Diet Diversity

Average household diet diversity consistently increased over time, showing sustained change. In fact, one-year post-intervention households on average consumed five or more food groups, out of the measured twelve food groups, during a 24-hour period (5.55 [1.3]), with increases in all food groups except a few (**Figure 1**.). Unlike diet diversity measurements for individuals, when measuring household diet diversity there is no standardized cutoff of food groups that will equal nutritional adequacy, thus a discussion of patterns is better suited for this data.^25^ At first glance, it appears the biggest increase is observed in the *Spices, Etc*. food group with an increase in over 80%, which could be interpreted as inconsequential as the gardens did not provide salt directly. However, upon further analysis, iodized table salt was the only spice reported, providing the essential micronutrient iodine, often lacking in the diets of poor rural populations globally.^26^ In addition, the consumption of many nutrient-dense foods increased by at least 20% from baseline to September 2019 to include foods in the following groups: *Cereals, Vegetables, Oils & Fats, Fruits*, and *Legumes, Nuts & Seeds*. Within the *Vegetables* food group, variety and nutrition density increased substantially, particularly with participants consuming more colorful varieties such as spinach, beets, carrots, and onions, providing health-promoting phytochemicals. They also increased the consumption of Vitamin A-rich vegetables such as dark leafy greens and carrots. However, since no biomarker data was collected on actual micronutrient levels in participants, no conclusions can be reported on actual nutrient status of participants or household members - a limitation that could be directed toward future study. However, the variety of vegetables offered by kitchen gardens likely contributed to household diet diversity both directly and indirectly which is consistent with other research in similar populations.^6,9,10^

Results also indicate that other foods groups, apart from vegetables, either continued to increase or were added one-year post-intervention. For example, consumption of foods in the *Cereals* food group in the form of sorghum, wheat, and maize flours or meals used for making porridge, all increased. These foods, grown primarily as cash crops in this region and purchased in the market, are considered some of the more expensive foods. This would suggest, that as households were able to save money from not purchasing vegetables from the market, they were left with more flexibility to spend money on other items such as cereals, salt, oil, and in a few cases milk and fish. Thus, it is possible that kitchen gardens not only increased the consumption of a variety of nutrient-dense vegetables, but also allowed for foods in other food groups to be consumed due to more income flexibility. Increasing income flexibility in a household is an indicator of progress toward poverty reduction by allowing for the purchase of nutrient-dense foods, rather than just those that meet energy needs.^6,27^ Therefore, further evaluation is warranted of the impacts of this intervention approach on income flexibility as a future research direction.

These changes in diet diversity did vary among the different cell groups. The reason is unclear but could be related to differences in the income source of the household among the groups. As reported, households whose main source of income was from working on the farms of others, saw lower diet diversity then those households that had someone working in the market or having formal employment (**Figure 4**.). This has also been reported in similar rural populations where those having a higher, more stable income source was associated with greater diet diversity.^6^ It is possible that the informal economy observed in rural areas where agriculture is the main provider, contributes to the seasonality and unpredictability of employment, representing a level of instability in this workforce versus those that have a steady and stable income.^27^ During the sunny season, many residents of Cyanika report no source of income due to the seasonality of crop production in the area. In accordance with historical data, this would account for the lower diet diversity previously reported during this time.^23^ However, as part of the intervention, rainwater collection tanks were distributed, and education on water management techniques was provided to help tackle this barrier. Concurrent with a study by Taruvinga et al. that examined barriers to increasing diet diversity in rural households located in South Africa, water management resources may have assisted in the success of the intervention by allowing for kitchen gardens to thrive and provide a diverse amount of foods to households through the sunny season.^6^ In the future, initiatives focused on innovative water management techniques for small-scale agriculture could greatly influence the success of nutrition-sensitive agriculture interventions as water is a constant barrier for many rural African communities.

### Stable Food Insecurity

The intervention did not result in significant changes in household food security. While two of the cell groups reported an increase in food insecurity after the intervention, most households maintained fairly consistent HHS scores reflecting moderate food insecurity as shown in Figure 7. Similar correlations with income source and household hunger were reported with diet diversity - those households whose main source of income came from working on the farms of others, saw greater food insecurity then those households that had someone working in the market or having formal employment (**Figure 4**.), suggesting that those with more stable employment also have more stable access to food.

While an explanation for these findings is not readily apparent, there are a few things to consider. First, through previous work with this population and community, the NGO has observed that the Rwandan culture in general, and especially in rural areas, exhibits an inclusive and compassionate collective attitude toward others. After the Rwandan genocide of 1994, the country as a whole adopted stronger cultural values such as unity, selflessness, volunteerism and humility which is apparent when one is immersed within the culture and exhibited by way of many of their government policies.^28,29,30^ This compassion is often translated to assisting others who are in need through the giving of money, food, household goods, employment opportunities and childcare. When considering this in the context of food security, many participants reported that they gave food away to others, possibly hindering their own household food security status.

Another potential factor influencing the household hunger score was insufficient training and resources for food preservation and storage as reflected during qualitative data collection discussed elsewhere. Food preservation practices and storage capabilities of extra food stores is a difficult barrier to overcome when refrigeration is nonexistent, and dry, reliable storage facilities are hard to maintain, as is the case in rural Rwanda. As reported in another study, this research corroborates this barrier also existing at the household level where inadequate storage facilities prohibited participants from keeping food for future periods, potentially contributing to the lack of improvement in food security.^31^ Therefore, more research is needed to develop ways that households can store their extra food while helping the community at the same time, so as not to degrade existing cultural values. Perhaps the next step for this community would be to consider larger community gardens that resonate with cultural values by potentially supplying more community members with food, and thus allowing those with kitchen gardens to store their excess for future use.

Regular border closures with neighboring Uganda, as well as the on-going Covid-19 pandemic, may have also impacted outcomes from the intervention. Cyanika, is located 4.6 kilometers from the Ugandan border where many residents cross daily for work and the trading of goods. As reported by study participants, when the border is closed, the amount of agricultural work available to residents of Cyanika is considerably less, causing many to go without an income for their household. These closures occur quite regularly due to violent and political conflict between the two countries, as well as regional health concerns. During this research, the border was closed several times for weeks to months. In addition, domestic stay-at-home orders due to the Covid-19 pandemic also affected household income sources. The source of household income in Cyanika was associated with variability in diversity and food security, thus it can be assumed that the disruption of these sources hindered any progress in food security. A recent study in Kenya and Uganda, reported decreases in diet quality and increased food insecurity during this time, specifically affecting income-poor households, corroborating with our observations.^32^ Although border closures and global health emergencies are not within the scope of control for this research, continued research on farmer resiliency and market integrity that informs policy is essential for the forward progress of nutrition-sensitive agriculture interventions as outlined by global organizations in recent years.^14,33^

### Limitations

The collaborative structure of the study within the community could result in response bias, as participants are also members of the community, and thus have a stake in the outcomes. This is especially true when the community and/or participants are receiving resources. In an attempt to mitigate this risk, triangulated data collection methods and participatory action research evaluation methods were employed. Second, we used household diet diversity scores as a proxy for evaluating dietary patterns and food access but did not collect the amount of foods consumed or nutrition-related biomarkers, both which measure nutritional adequacy. Therefore, we cannot reliably predict that the foods consumed were in sufficient amounts to meet nutritional adequacy. Third, the exploratory nature of this study with a small sample size and lacking a control group, provided results that can only suggest the possibility that the intervention had a positive impact on household diet diversity, and thus subsequent dietary patterns. Our intention was to provide direction for future research and a potential intervention structure that could yield sustained changes to dietary patterns and food access.

## Conclusion

This study shows that collaborative community-based nutrition-sensitive agricultural interventions in rural low-income Rwandan populations, have the potential to increase household diet diversity to encourage sustained change in dietary patterns for nutritional adequacy. Using kitchen gardens as the conduit for change, households can increase their consumption of home-grown vegetables, as well as other foods, by increasing household income flexibility. These findings are in accordance with other studies in similar sub-Saharan African populations, and thus the intervention approach could translate to similar populations. Reasons for the lack of improvements in food insecurity in the face of increased dietary diversity are unclear but suggest that additional research is necessary regarding the systems that affect food availability and agricultural markets needed to enact changes in food security.

## Data Availability

All shareable data is included in the submitted manuscript. Other data is associated with personal identifying information therefore protected under the Colorado State University Institutional Review Board.

## Acknowledgements

Authors would like to thank Village Makeover, Rotary International, The Department of International Programs at Colorado State University, Administrative Secretary of Cyanika, Rwanda Project Managers Jean Amie Hasani and Jacques Ngirahahizi, and study participants.

